# High-resolution 3-D imaging for precise staging in malignant melanoma

**DOI:** 10.1101/2020.07.22.20159103

**Authors:** Simon F. Merz, Philipp Jansen, Ricarda Ulankiewicz, Lea Bornemann, Tobias Schimming, Klaus Griewank, Zülal Cibir, Andreas Kraus, Ingo Stoffels, Timo Aspelmeier, Sven Brandau, Dirk Schadendorf, Eva Hadaschik, Gernot Ebel, Matthias Gunzer, Joachim Klode

## Abstract

High-resolution imaging of sentinel lymph nodes (SLN) from melanoma patients is a crucial approach to specify staging and determine individuals requiring adjuvant treatment. Current histologic SLN analysis has the substantial drawback that only a small portion of the node is sampled while most of the tissue is discarded which might explain the high false-negative rate of SLN diagnosis. Therefore, we developed an algorithm-enhanced light sheet fluorescence microscopy (LSFM) approach to three-dimensionally reconstruct the entire SLN with the power to identify single tumor cells. We comprehensively quantified total tumor volume while simultaneously visualizing cellular and anatomical hallmarks of the associated SLN architecture. In a first-in-human prospective study (21 SLN from 11 melanoma patients), LSFM not only identified all metastases seen histologically, but additionally detected metastases not recognized by routine histology. Thus, our 3-D digital pathology approach can increase sensitivity and accuracy of SLN-metastasis detection and potentially alleviate the need for conventional histopathological assessment in the future.

## Introduction

According to the World Health Organization, the incidence of melanoma is increasing faster than any other major cancer worldwide. In the United States, melanoma is the fifth most common cancer, posing a substantial health and economic burden (Livingstone et al., 2011). Status assessment of draining sentinel lymph nodes (SLN) is an essential diagnostic procedure incorporated in clinical management guidelines of melanoma and other cancers, since SLN tumor burden is the most important prognostic factor for overall survival (Gershenwald et al., 2017b; Resch and Langner, 2013; Sharma et al., 2017; Zahoor et al., 2017). Prognosis further correlates with several characteristics such as size and location of metastases within SLN, cumulatively identifying patients in need of adjuvant treatment (van Akkooi et al., 2006; van Akkooi et al., 2008; van Akkooi et al., 2009; van der Ploeg et al., 2011). However, SLN positivity rates vary widely depending on applied histopathological procedures (Riber-Hansen et al., 2012; Riber-Hansen et al., 2008; Spanknebel et al., 2005).

Conventional histology of two-dimensional (2-D) tissue sections is the currently used gold standard for SLN staging (Cook et al., 2019). However, by design this approach assesses only a few representative slices of the node, while discarding the majority of the tissue (Riber-Hansen et al., 2012) thus potentially explaining the reported high false-negative rates of SLN diagnosis (up to 38%), (Nieweg, 2009) today.

Despite this knowledge, currently no histological approach exists that would allow analyzing the entire SLN volume in a routine setting. While complete sectioning/analysis of the entire SLN is theoretically possible, the time and cost involved prohibit routine application. However, not reliably detecting single or isolated tumor cells in the SLN is a considerable short-coming of current histology protocols, as recent data demonstrate that even very small deposits affect long-term prognosis of melanoma patients (Murali et al., 2012; Scheri et al., 2007; van der Ploeg et al., 2014). In fact, determining the exact size of metastatic deposits is critical as melanoma-specific survival is reduced with increasing SLN tumor burden (Gershenwald et al., 2017a) and microscopic SLN tumor load is a stratification factor for adjuvant treatment (Eggermont et al., 2016; Long et al., 2017). Collectively, the reliable detection of few melanoma cells within the three-dimensional (3-D) space of entire SLNs with simultaneous volume quantification and 3-D localization of tumor foci relative to anatomical landmark structures within the SLN remains a diagnostically relevant challenge not adequately addressed to date (Cook et al., 2019; van Akkooi et al., 2009). These considerations highlight the unmet need for a method capable of determining exact tumor volume and location within a SLN.

We and others recently developed clearing techniques to generate optically transparent samples from many different soft and hard tissues of experimental animals (Kirst et al., 2020; Klingberg et al., 2017; Masselink et al., 2019) as well as of human organs (Belle et al., 2017; Zhao et al., 2020) with subsequent rapid 3-D reconstruction at cellular resolution using light-sheet fluorescence microscopy (LSFM) (Grüneboom et al., 2019a; Grüneboom et al., 2019b; Männ et al., 2016). Recent advances enable the imaging of strongly autofluorescent tissue (Merz et al., 2019), a highly relevant improvement for skin draining SLN containing pigmented cells. So far, this was an unsolved problem in assessing diagnostically relevant pigmented human samples (Nojima et al., 2017). Since organ clearing combined with LSFM can provide 3-D information of large tissue samples while being able to identify single cells, it might improve SLN staging. However, this had not been applied so far in a clinical setting.

Here, we present a new method making the entire volume of human SLN amenable to 3-D inspection at cellular resolution using optical clearing and immunofluorescence-based LSFM. Our workflow enables whole-mount staining and subsequent identification of anatomical hallmarks of SLN (capsule, vascularization, immune cells) with embedded pigmented melanoma cells. We demonstrate that algorithm-enhanced LSFM provides a browsable histo-like optical slice library for fast and reliable SLN status determination and tumor environment assessment. All data were acquired in a cross-sectional study comparing our new LSFM imaging approach with gold-standard clinical histopathological analysis, with the goal of assessing the performance relative to the current standard of care and potentially suggesting translation of this new imaging approach to optimized diagnoses of melanoma. The results demonstrate that LSFM was able to identify 100% of metastases detected by standard histopathology, yet it also discovered small tumor foci, that had initially been overlooked. This directly led to changes in clinical management of an affected patient, thus providing evidence, that 3-D LSFM has the potential to fundamentally improve SLN staging in melanoma.

## Results

### Establishment of a staining panel for 3-D SLN analysis

For 3-D cellular LSFM-based analysis, human lymph nodes (LN) were optically cleared according to previously described ethyl cinnamate-based protocols (Klingberg et al., 2017; Merz et al., 2019). A peroxide bleaching step was used to attenuate endogenous autofluorescence and destroy pigments (especially hemoglobin and melanin) that otherwise reduce imaging quality. Tissue autofluorescence alone was sufficient to visualize anatomical structures, like the LN capsule or blood vessels (Figure 1a), as well as to 3-D reconstruct an entire LN, revealing a complex surface architecture (Figure 1b, Supplementary Video 1). However, due to the intra-image variance of signal brightness of e.g. fat (dark), LN stroma (medium bright) and extremely bright erythrocytes, simultaneous display of all entities in a single image was impossible. Therefore, we developed RAYhance, a multiscale contrast compression algorithm, that allows display of LSFM data as a browsable histo-like stack, like 2-D formalin-fixed paraffin-embedded (FFPE) sections (Figure 1a, Supplementary Figure 1).

**Figure 1:**
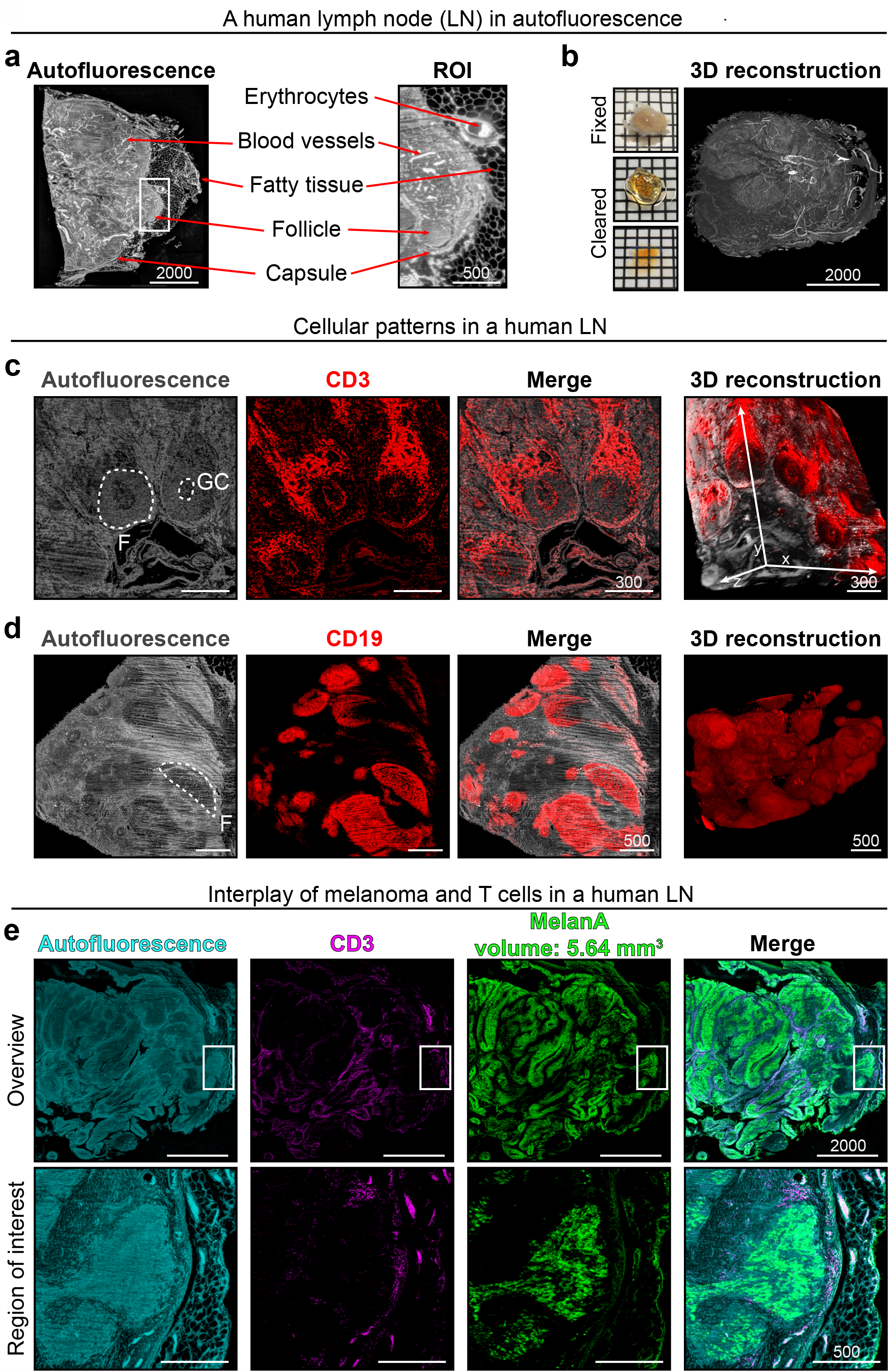
Anatomical and cellular patterns in a human lymph node (LN). (**a**) Anatomical features of a human LN including position of the capsule, blood vessels or fatty tissue can be derived from the tissue’s autofluorescence signal alone. These features are highlighted (red arrows) in the algorithm-enhanced optical LSFM overview slice (left) and the region of interest (ROI, white rectangle; also see Supplementary Figure 1). (**b**) Macroscopic images and a 3-D reconstruction of the same human LN based on autofluorescence (Supplementary Video 1). In order to reveal cellular patterns within a human LN, we applied whole-mount staining prior to clearing. Again, using autofluorescence (gray) alone, follicles (F) and even germinal centers (GC) could be identified. This was confirmed by visualizing anti-CD3 staining (T cells, **c**) and anti-CD19 staining (B cells, **d**). On the right side of each row of LSFM optical slices is a 3-D reconstruction of the data set. (**e**) A melanoma metastasis in a human lymph node revealed by anti-MelanA staining (green). Anatomical and cellular patterns can be assessed using autofluorescence (cyan) and T cells (magenta), respectively. Region of interest (bottom row) shows MelanA positive cells surrounded by T cells in close proximity to the capsule (Supplementary Video 2). Scale bar values in µm, squares in macroscopic images 2×2 mm. All optical slices are RAYhance-processed.

#### BOX to explain purpose of RAYhance

The grey value dynamics of LSFM images are around 10k – for a given ambient brightness the observer’s visual system however is able to differentiate only around 100 to 200 different grey values ranging from what is perceived as black to white. Therefore, the standard approach is to employ a visualization tool’s ability to shift the recognizable dynamic range to a squeezed min/max-intensity range of the raw image, the so-called grey scale compression. Employing grey scale compression inevitably introduces severe under- and overexposure, additional gamma settings alleviate this to a certain extent by introducing a non-linear transfer function. The purpose of RAYhance is to extract all structures hidden in the original raw data so that they could be detected by a human observer at one glimpse without any further visualizing tool interactions. This means that subtle structures of vastly dim regions shall be visualized in the same image together with structures of very high exposed regions. To reach this goal RAYhance firstly calculates different image resolutions of contrast versions of the original intensity image, which is a multiscale approach. Secondly, the multiscale contrast versions are non-linearly transformed in order to enhance very low contrasts and to decrease very high contrasts which relate to structures that are perceivable anyhow. Thirdly, it recombines them into one resulting artificial image of the raw image’s original resolution. It appears as a modified intensity image which delivers all characteristics formulated in RAYhance’s purpose.

To visualize prominent intra-nodal cellular patterns, we adapted and modified previously described whole-mount staining procedures (Belle et al., 2017; Renier et al., 2014). CD3-expression revealed the T cell zone and germinal centers within the presumed autofluorescence-dim B cell follicles (Figure 1c). Here, RAYhance was instrumental to simultaneously visualize thousands of clustered T cells (strong signal) in the T cell zone and single cells (dim signal) in the B cell follicles or germinal centers (Supplementary Figure 2). CD19-expression confirmed B cell enrichment in the autofluorescence-dim and T cell poor regions (Figure 1d). In summary, autofluorescence combined with fluorescent immune cell staining portrayed the overall anatomical structure and major cellular distribution patterns of human LNs (Figure 1).

Next, we used anti-MelanA staining, a routinely employed melanocytic marker (Weinstein et al., 2014), to visualize melanoma metastases. This approach enabled 3-D quantification of the total volume of MelanA positive structures and simultaneously demonstrated the interplay of immune cells with MelanA-expressing elements (Figure 1e). Again, RAYhance processing permitted the display of single CD3 and MelanA positive cells (Supplementary Video 2, 3).

### LSFM analysis is compatible with subsequent routine histology

To exclude impact of peroxide-bleaching, whole-mount staining, clearing and imaging on the histopathological appearance of human tissue and melanoma markers, we took biopsies of the same sample after simple fixation (routine protocol), bleaching and LSFM imaging. Routine FFPE stainings and immunohistochemistry (IHC) were performed on all samples. Importantly, no protocol-associated discernible changes in cellular morphology (hematoxylin/eosin staining) or staining quality and distribution of the histopathologically relevant melanoma markers MelanA and S100 in clinical grade FFPE sections were detected (Supplementary Figure 3). Thus, our approach allows an intra-sample comparison of findings in both, LSFM and subsequent gold standard FFPE analysis.

### Integrating LSFM into the routine SLN analysis of melanoma patients

Utilizing this, we incorporated our approach into the ongoing routine SLN analysis of melanoma patients to benchmark the performance of the protocol and to test the sensitivity and specificity of LSFM for the detection of micrometastases. According to the current gold standard, SLN are post-surgically processed for FFPE histopathological analysis of individual representative tissue slices. Hence, resected SLN were first analyzed by LSFM and then routinely processed for ensuing histopathology (Figure 2). During routine slicing of paraffin blocks, gap sections (300 µm – 600 µm) between the relevant FFPE tissue slices are typically discarded and no longer available for later analysis. In contrast, all LSFM samples, once paraffinized, were sequentially cut and non-relevant slices were stored allowing correlation of all possible unique LSFM findings with follow-up histology (Figure 2).

**Figure 2:**
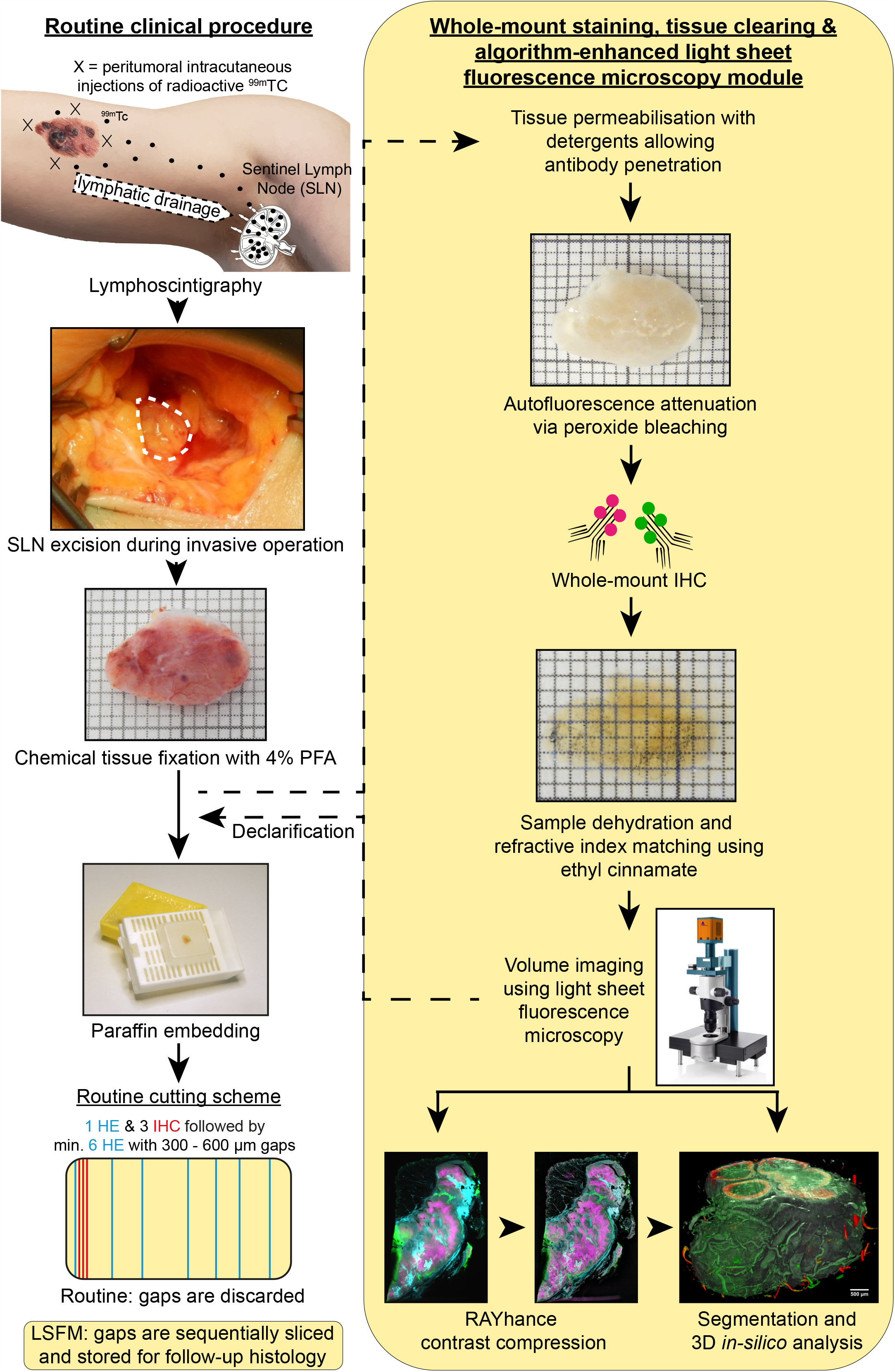
Light sheet fluorescence microscopy (LSFM) in melanoma diagnostics. Incorporation of LSFM analysis (right yellow column) into the routine clinical diagnostics process to analyze human sentinel lymph nodes of melanoma patients (left column). Lymphoscintigraphy with radioactive technetium (^99m^Tc) allows the determination of the local draining lymph node (sentinel lymph node, SLN) of the primary melanoma area. The SLN is excised and post-surgically fixed using formaldehyde (4% PFA). In the fixed state, the SLN is routinely paraffin embedded and sliced for hematoxylin/eosin (HE) staining and immunohistochemistry (IHC). Here, only few representative slices are analyzed for melanoma metastases per sample. To analyze the whole SLN tissue, LSFM analysis was executed after sample fixation with 4% PFA and before performing gold standard histopathological analysis. The SLN was prepared for LSFM analysis using tissue permeabilization, autofluorescence attenuation, whole-mount IHC and tissue clearing by ethyl cinnamate (ECi). Obtained volumetric LSFM data can be algorithm-enhanced and 3-D-segmented for exact volume quantification. After imaging, samples were declarified and transferred back to routine processing. Note that lymph node tissue between the relevant paraffin slices for histopathological analysis (gap sections) are routinely discarded and cannot be considered for SLN status determination. However, in order to validate LSFM findings throughout the entire sample, gap sections were sequentially sliced and stored to allow for follow-up formalin-fixed paraffin-embedded (FFPE) analysis. Squares in the macroscopic LN images are 1×1 mm.

With the staining procedure and panel established, we implemented a prospective study assessing the metastatic status of SLN. In total, 21 SLNs of 11 melanoma patients (Supplementary Table 1) were analyzed without prior selection of their location or size (Supplementary Table 2). We cut several SLN into smaller pieces to reduce the processing time before subsequent obligatory histopathological analysis and to minimize the delay of patient diagnosis. Furthermore, this ensured robust and complete whole mount staining within an acceptable time frame. We determined that LSFM sensitivity compared to the gold standard in 105 SLN-derived samples was 100% and the false negative rate was zero (Supplementary Table 2). Importantly, LSFM identified metastases in one patient that were not found in the initial routine histopathological assessment (see below).

### Diagnosis of a capsular nevus in LSFM and histology

In a 1.7 × 1 cm large inguinal LN (patient #4), blood residues and pigmentation were successfully bleached before LSFM analysis (Figure 3a). Three pieces of the node were found positive for intra-capsular MelanA positive cells and hence suspected to contain a metastasis (Figure 3b,c). LSFM overview scanning revealed MelanA positive signals within the LN, while T cell structures remained unperturbed (Figure 3b, Supplementary Video 4). However, some MelanA staining artifacts were also encountered. Most prominent was an unspecific accumulation of anti-MelanA signal at sites of physical manipulation, that did not correlate with follow-up FFPE IHC (cutting-edge artifact, CEA, Supplementary Figure 4). In contrast, a discernible MelanA background staining was observed by both LSFM and FFPE in fatty tissue (fat artifact, FA in Figure 3b). Higher magnification LSFM of regions of interest (ROI) showed that i) the localization of the MelanA positive cells was intracapsular, ii) the spread of MelanA positive cells was filamentous and heterogeneous and iii) the associated follicular structure of the LN remained intact (Figure 3c,d). 3-D segmentation of the total tissue block (96.8 mm^3^), the LN capsule used for assessment of the LN tissue volume (70.7 mm^3^), the CD3 (35.9 mm^3^) and the intracapsular anti-MelanA (0.24 mm^3^) signal allowed volume quantification. Moreover, MelanA positive cells were visualized as a sparsely growing intracapsular network, sheathing the round follicular structures (Figure 3c,d, Supplementary Video 5). Additionally, using autofluorescence, we determined the sinus volume of the LN tissue to be 14.5 mm^3^. Furthermore, CD3 staining allowed the quantification of cellular and anatomical hallmarks like follicles and germinal centers. In the tissue piece shown in Figure 3c, we counted 161 follicles with 154 germinal centers, occupying a median volume ± interquartile range (IQR) of 1.7×10^7^ ± 2.1×10^7^ µm^3^ and 2.8×10^6^ ± 4.9×10^6^ µm^3^, respectively. 124 (77%) of the follicles were secondary follicles, of which 19 (15.3%) contained more than one and up to three germinal centers. Only 37 (23%) were primary follicles. Follicles also differed significantly (p<0.0001, Mann-Whitney *U*-test) in volume with 5.2×10^6^ ± 6.0×10^6^ µm^3^ for primary and 2.2×10^7^ ± 2.2×10^7^ µm^3^ for secondary follicles (Supplementary Video 6). In total, 7% of the cortex/paracortex tissue (excluding the medulla) consisted of follicles and 17.5% of this space was taken up by germinal centers, demonstrating the power of LSFM to precisely quantify LN functional zones.

**Figure 3:**
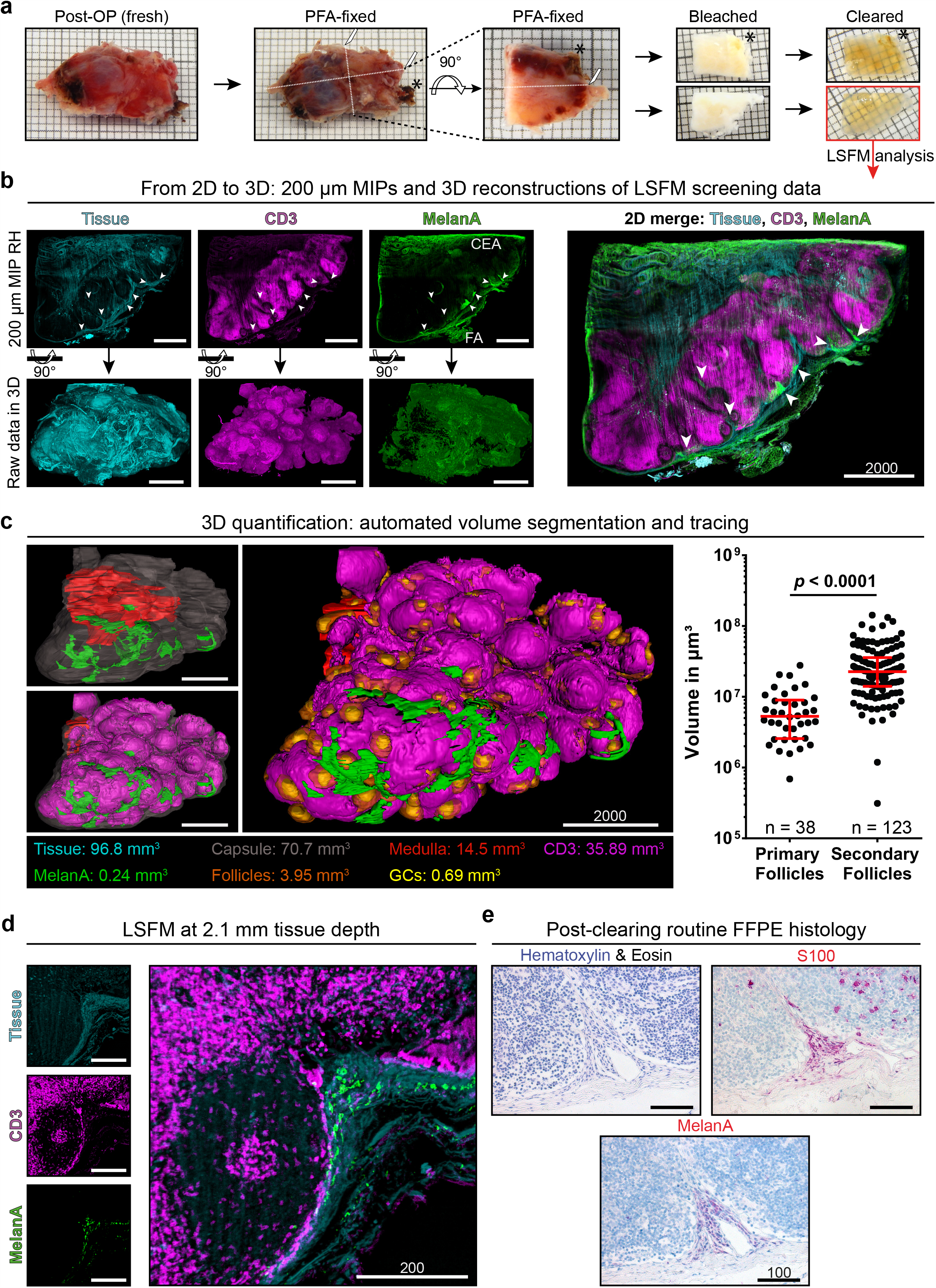
From 2-D to 3-D and back – a capsular nevus in all protocol stages. (**a**) Macroscopic images of an inguinal human sentinel lymph node (patient #4, S8) at various steps of the protocol. Due to size limitations the sample was cut multiple times before commencing with the protocol. Dashed lines with a scalpel symbol indicate incisions performed in conformity with the gold standard processing protocol. The black asterisk highlights a pigment-rich region, allowing tracking after bleaching and clearing (in ECi). The lower sample in the clearing panel is shown throughout the analysis process, which is indicated by the red box and arrow downwards. One square equals 1×1 mm. (**b**) Upper panel shows 200 µm maximum intensity projections (MIP) of algorithm-enhanced (RH = RAYhance) fluorescence signal of general tissue autofluorescence (cyan), anti-CD3 (T cells, magenta) and anti-MelanA (green). Clusters of intracapsular MelanA signal are highlighted by white arrows. Note that anti-MelanA staining exhibits artifacts, e.g. cutting-edge artifact (CEA) and fat artifact (FA), which makes rendering the raw signal difficult (Supplementary Figure 4). The lower panel shows the corresponding 3-D reconstructions for every channel (Supplementary Video 4). On the right, a 2-D MIP color merge is depicted. (**c**) 3-D segmentation and quantification of LN tissue within the capsule (gray), T cells (magenta), medulla (red), follicles (orange), germinal centers (GCs, yellow) and MelanA signal (green) within the capsule (Supplementary Video 5, 6). The right scatter plot displays the size distribution of quantified primary follicles (n = 37), without GCs, and secondary follicles (n = 124), containing one or more GCs, in the shown sample (biological n = 1). The median volume ± interquartile range displayed in red is 5.2×10^6^ ± 6.0×10^6^ µm^3^ for primary and 2.2×10^7^ ± 2.2×10^7^ µm^3^ for secondary follicles. *p*-value (*p* < 0.0001) was calculated using a two-tailed Mann-Whitney *U*-test. (**d**) One optical slice imaged at 2.1 mm tissue depth with 12.6x magnification. The intracapsular position of the elongated MelanA positive cells is clearly visible. (**e**) Diagnostically relevant FFPE sections after declarification, paraffination, physical sectioning and routine staining. Anti-S100 and anti-MelanA antibody signals are visualized in red with a blue nuclear counterstain (Hematoxylin). All scale bar values are given in µm, all optical slices are RAYhance-processed.

Subsequent routine histopathological assessment (Figure 3e) confirmed the MelanA-expressing cells to be i) intracapsular, ii) of elongated shape with inconspicuous nuclei, iii) next to MelanA being also positive for the melanocytic marker S100. Based on these findings, a benign capsular nevus was diagnosed. This sequence of events showed that LSFM revealed all necessary information required to allow a diagnosis, but also highlights the importance of compatibility and close interplay between 3-D LSFM and routine histology.

### Identification of melanoma metastases

In another axillar SLN (patient #9), five of twelve pieces were found to be positive for MelanA (Figure 4). The signal was primarily enriched directly below the capsule but also extended into the parenchyma. At the borders of the compact, interconnected cell mass, multiple single cells and multi-cellular clusters could be identified, like a spray within the LN tissue (Figure 4b, Supplementary Video 7). It was now possible to 3-D quantify the total MelanA positive infiltrate. MelanA positive cells together (in all tissue pieces) occupied a total volume of 5.35 mm^3^. With respect to the whole lymph node volume of 503 mm^3^ (excluding all extracapsular fat and tissue), we determined that 1.06% of the entire SLN was enriched with MelanA positive cells (Figure 4), information not obtainable from clinical histopathology. Importantly, the distribution patterns of MelanA positive cells observed in LSFM correlated precisely with subsequent histopathological stainings (Figure 4c).

**Figure 4:**
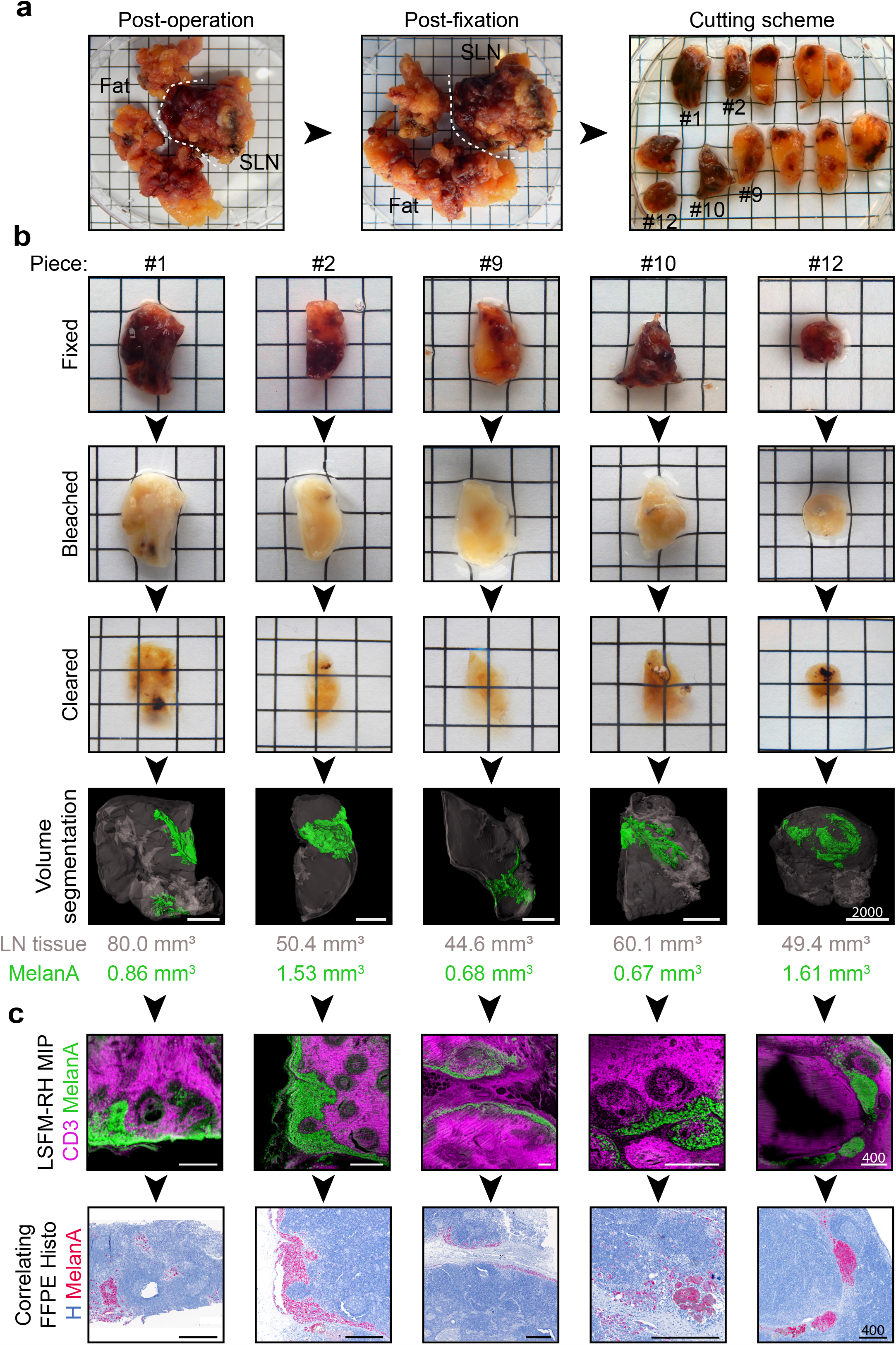
Melanoma metastasis assessment in human SLNs using LSFM. (**a**) Macroscopic image of an axillar human SLN (patient #9, S18) after operation and fixation. The surrounding fat is cut and processed separately for routine histology. The SLN is cut into twelve smaller pieces, five of which are found to be metastasized. (**b**) *From top:* Panels of macroscopic images showing the five metastasized SLN pieces, each one in a column, after fixation, bleaching and clearing (in ECi). Some residual pigmentation can be seen macroscopically. Impact on LSFM imaging see Supplementary Figure 5 and Supplementary Video 8. The LN tissue within the capsule (gray, without surrounding fat) as well as the subcapsular MelanA signal (green) were volume segmented and quantified (Supplementary Video 7). The total LN tissue volume of all twelve pieces combined amounted to 503 mm^3^, the total MelanA volume to 5.35 mm^3^ (1.06%) (**c**) *Top:* LSFM MIPs of three subsequent RAYhanced (RH) optical slices, showing anti-MelanA signal (green) and anti-CD3 signal (magenta). *Bottom:* Correlating FFPE slices of follow-up anti-MelanA histology (MelanA = red, Hematoxylin = blue). Squares in macroscopic images are 5×5 mm, scale bar values in µm.

Despite performing a peroxide bleaching step, residual pigmentation was present in some tissue pieces. This impacted LSFM imaging with shadow artifacts like stripes or blurriness (Supplementary Note 1). However, despite heavy pigmentation of e.g. piece twelve (Figure 4b), we were able to completely 3-D quantify the anti-MelanA signal, highlighting the importance of using infrared laser light (785 nm excitation) for tumor detection. In this LN piece, the association of MelanA positive cells with a major blood vessel was particularly striking (Supplementary Figure 5, Supplementary Video 8). Correlative follow-up histology revealed that pigmentation was sometimes, but not always, co-localized with MelanA positivity (Supplementary Figure 5).

### Discovery of hidden metastases

In another inguinal SLN (patient #11) with a total intracapsular volume of 749.3 mm^3^, we determined MelanA positive cells with a total volume of 0.03 mm^3^ (0.004%) in 1 of 11 pieces. This volume consisted of two intranodal volumes of 0.013 mm^3^ and 0.006 mm^3^ and an extracapsular but intravascular volume of 0.011 mm^3^. Each site alone was much larger than the 0.1 mm diameter (0.0005 mm^3^ as a sphere) measurement considered diagnostically relevant (van Akkooi et al., 2009; van der Ploeg et al., 2014). However, the subsequent routine FFPE histology analysis did not identify a SLN metastasis. Neither MelanA nor S100 positivity were observed within the SLN of the sections analyzed. The SLN was therefore declared “negative” according to the gold standard histopathological analysis. To further investigate the MelanA positive signals only observed in LSFM, we performed an additional follow-up FFPE histology on preserved gap sections (Figure 2) of the SLN tissue. Here, we identified MelanA positive cell clusters confirming the previous LSFM observations (Figure 5, Supplementary Video 9). As first discovered in the LSFM analysis, small morphologically atypical cells penetrated the lymph node within an afferent lymph vessel. Those cells were distributed within the subcapsular sinus and showed initial infiltration of the parenchyma. Based on the morphology, the cellular distribution and the histopathological staining, this cell accumulation was recognized as a melanoma metastasis. The status of the SLN was subsequently re-evaluated and changed to “positive”. Hence, LSFM-guided histopathological analysis enabled identification of a melanoma metastasis in a SLN that would have been classified as “negative” if assessed by conventional histopathology protocols alone. Based on these results, the patient was reassigned to the interdisciplinary melanoma tumor board and was subsequently offered access to adjuvant checkpoint blocker therapy.

**Figure 5:**
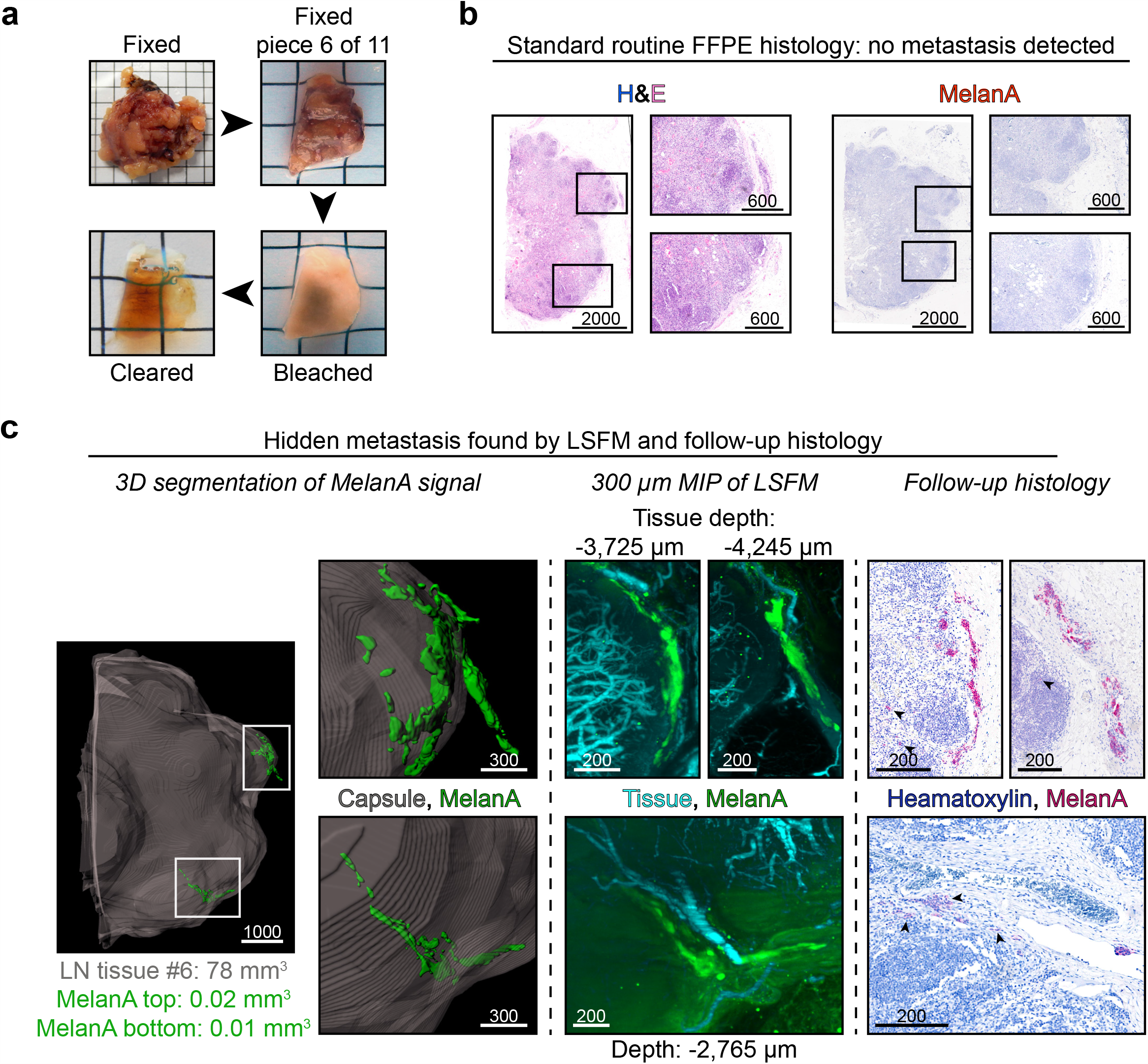
LSFM reveals metastasis missed by the gold standard procedure. (a) Macroscopic images of an inguinal human SLN (patient #11, S21) after operation and piece number six after fixation, bleaching and clearing (in ECi). (**b**) Routine FFPE histology found no MelanA positive cells within the lymphnode in piece six, nor any other piece of the LN (not shown). Enlargements (black rectangles in overview) correspond to regions of interest from (c). (**c**) MelanA signal could be detected at two sites (white rectangles) of tissue piece six. *Left:* Volume segmentation of MelanA signal (green) and the capsule allowed 3-D quantification. Enlargements of upper and lower MelanA events can be correlated with both LSFM MIPs (middle) and LSFM-guided follow-up histology (right). *Middle:* 300 µm maximum intensity projections (MIPs) of tissue autofluorescence (cyan) and MelanA signal (green). The MelanA event in the top panel consists of two parts, in total stretching over more than 300 µm in Z. The left part is located in the sub-capsular sinus (Supplementary Video 9). The right part is outside of the capsule, located within an afferent lymph vessel (see histology). The MelanA event in the lower panel is closely associated with a blood vessel. *Right:* Additional LSFM-guided follow-up FFPE IHC stainings against MelanA (red) confirmed LSFM findings. Black arrows highlight MelanA positive cells that are easy to miss. In the upper left image, singular parenchymal cells can be identified. Squares in macroscopic images are 5×5 mm, scale bar values in µm.

## Discussion

Over the recent years, several studies demonstrated great efficacy of immune checkpoint inhibitors and MAP-kinase directed drugs in advanced melanoma. These novel treatments are currently approved for adjuvant treatment in stage III disease (Amaria et al., 2019; Grob et al., 2018). The current standard determining stage III patients is the histopathological analysis of the SLN. Therefore, the SLN status is not only the most important prognostic factor in melanoma but also SLN tumor burden is the most important stratification factor refining subgroups for adjuvant treatment, hence for prolonging melanoma survival. Currently, the SLN tumor burden is measured as the diameter of the largest lesion detected in histopathological analysis. While proven effective and widely used for decades, it has limitations. As established protocols merely analyze representative slices (Cochran et al., 2000; Mitteldorf et al., 2009; Riber-Hansen et al., 2012; Satzger et al., 2007; Starz et al., 2001), only a very small portion of the SLN is sampled for histology. Without prior knowledge of tumor cell localization, the SLN is sectioned according to a predetermined cutting scheme (Figure 2, (Cook et al., 2019)). This black-box approach is prone to error allowing micrometastases to be missed entirely. When discussing the detection of total melanoma volume in SLN, several considerations are important. First, the smallest tumor considered diagnostically relevant in FFPE diagnostics has a diameter of 0.1 mm (van Akkooi et al., 2009; van der Ploeg et al., 2014). Assuming an average melanoma cell size of ∼20 µm diameter and a spherical volume, such a tumor would contain ∼125 cells in 3-D. Second, a small SLN (1 cm diameter) has a volume of ∼5.2*10^11^ µm^3^ while the tumor cell volume would be ∼4.2*10^3^ µm^3^. Thus, even a small SLN offers ∼10^8^ different positions for a single melanocyte or ∼10^6^ sites for minimally relevant tumor deposits to be located. Given that melanocytic metastases differ in cellular form (e.g. ellipsoid or irregular), a complete sequential sectioning of the SLN in 5 µm steps would be mandatory to guarantee not missing a metastasis by conventional histology. As multiple stainings of a single melanoma metastasis are required (e.g. H&E, anti-MelanA, anti-S100), a melanoma cell may be missed if not present in at least two sequential sections. Hence, ∼2,000 serial sections would be necessary to definitively declare a 1 cm SLN positive or negative. As SLN are often larger, the number of required sections would frequently be substantially higher. Thus, a complete histological analysis of a SLN is not feasible in a routine setting.

To overcome this problem and optimize diagnosis and treatment in melanoma, we present a 3-D digital LSFM pathology approach to analyze the whole SLN and to identify single, marker-positive cells in their anatomical context. To our knowledge, this study is the first to investigate the potential benefit of LSFM-based determination of SLN status compared with conventional histopathology. The results of our ongoing study already demonstrate that this approach allows the complete 3-D tumor assessment, additionally showing melanocyte interactions with the anatomical and cellular environmental cues with a reasonable investment of time and expenses. The precise 3-D localization of metastases with respect to anatomical hallmarks (e.g. blood/lymph vessels, immune cells, stroma) could yield important new insights into metastasis formation and biology, possibly impacting prognostic assessment and treatment in the future. Using machine learning or spot detection algorithms it might even be possible to predict which patient could potentially benefit from a checkpoint inhibitor therapy by quantifying T cell infiltration in SLN metastasis (Tang et al., 2016).

In comparison to routine FFPE histology, which can theoretically be available for histopathological assessment within 2-3 days, our LSFM approach currently still requires 12 additional days until data acquisition is complete (Supplementary Table 3). Although the direct comparison of time required by FFPE-histology analyzing single representative slices with LSFM acquiring hundreds of slices per sample is difficult, an important future challenge is to reduce protocol times. This could be achieved by shortening tissue penetration time of staining molecules by e.g. electrostatic fields (Lee et al., 2016), microwave enhanced diffusion (Katoh, 2016; Katoh et al., 2009) or smaller staining molecules like nanobodies (Wang et al., 2016) that allow considerably faster diffusion (Gefen et al., 2008). Furthermore, the development of both, artifact-free antibodies and strong, photobleaching-resistant infrared fluorophores compatible with whole-mount staining, clearing, LSFM and follow-up histology are of high importance. That antibodies can be applied without causing artifacts is demonstrated by the anti-CD3 or anti-CD19 stainings (Figure 3). The benefits of infrared dyes in whole organ LSFM were demonstrated in a tumor measured despite the illumination and emission shadow of residual pigmentation (Supplementary Figure 5, Supplementary Video 8), a significant improvement of previous reports investigating human LN (Nojima et al., 2017).

Another problem connected to LSFM (and any in-depth imaging approach) is the size of the resulting data sets that need to be processed and stored (Poola et al., 2019). Importantly, significant information contained in raw LSFM images can also be obscured when using conventional and strictly linear digital display options for visual inspection that inevitably lead to display saturation in maximal brightness and/or darkness. Therefore, we introduced RAYhance as a multiscale contrast compression algorithm that obeys the characteristics of the human visual system and processes the primary image in several multiscale contrast images. This allows the simultaneous representation of all clinically relevant image entities even if they stem from formerly disparate intensity regions. However, RAYhance, in its current version, being optimally suited to adjust the autofluorescence channel can only be a starting point. Other signals such as the tumor may need more delicate adjustments to fully represent the biological distribution without creating false positives. New approaches such as deep learning may recognize disease-defining features in LSFM data, not discernible to the human eye, yet bearing substantial diagnostic power (Belthangady and Royer, 2019).

Currently, our three channel setup already allows identification of at least seven distinct entities of SLN architecture such as medulla, follicles and germinal centers (Figure 3). To further improve landmark identification, future developments should overcome current limitations of three color LSFM. Using more infrared laser lines and more fluorescent dyes between 600-850 nm emission will allow multiplexed analyses. In conjunction with the advent of process automatization, our approach could potentially represent the starting point of a fast, routine stand-alone method of multicolor 3-D tumor analysis in human tissues. It is tempting to speculate that, in the future, pathologists might browse through 3-D data sets of whole tissues in search of conspicuous structures (Poola et al., 2019) rather than spend hours behind microscope oculars examining thin sections.

The diagnostic potential of the presented approach is strengthened by the fact that it not only identified all metastases seen histologically, but additionally detected two metastases not recognized by routine histology. Based on this finding, the interdisciplinary tumor board changed their recommendations and this patient was offered an adjuvant therapy with an immune checkpoint inhibitor. Therefore, our novel technique already provides a further stratification factor for adjuvant treatment potentially prolonging melanoma-specific survival.

Importantly, the application of LSFM-guided histology in diagnostics is not restricted to melanoma. Apart from analyzing SLN of other cancer types (Nojima et al., 2017), the procedure can also be applied to other diagnostically relevant tissue samples (Merz et al., 2019; Nojima et al., 2017; Poola et al., 2019). In conclusion, our workflow can guide and potentially replace FFPE histopathology as a novel gold standard for SLN staging.

## Methods

### Lead contact and materials availability

This study did not generate new unique reagents. Further information and requests for resources and reagents should be directed to and will be fulfilled by the Lead Contact, Joachim Klode.

### Experimental model and subject details

#### Clinical trial

This prospective study was approved by the local ethics committee of the Medical Faculty, University Hospital Essen, Germany (17-7420-BO) and registered at the German Clinical Trials Register (DRKS00015737) with the Universal Trial Number (UTN) U1111-1222-2725 of the World Health Organization (WHO). Written, informed consent was obtained from all patients before entering the study.

#### Inclusion and exclusion criteria (human cohorts)

Eleven patients fulfilled the following inclusion criteria: age >18 years, histologically confirmed diagnosis of malignant melanoma with stage Ib or II (tumor depth of ≥1.0 mm) scheduled to undergo sentinel lymph node excision (SLNE) according to American Joint Committee on Cancer (AJCC 2009). Characteristics of included patients are listed in Supplementary Table 1 and obtained sample characteristics can be found in Supplementary Table 2.

#### Sentinel lymph node excision

SLNE is performed as a standard procedure at the Department of Dermatology, University Hospital Essen, Germany, according to the guidelines of the German Association of Dermatology (Deutsche Dermatologische Gesellschaft, DDG) (Garbe et al., 2008). Prior to SLNE, lymphoscintigraphy using radioactive technetium and SPECT/CT (Stoffels et al., 2012) was performed and subsequent SLNE was performed either under tumescent local anesthesia or general anesthesia as previously described (Stoffels et al., 2019). Preparation and subsequent excision of all marked lymph nodes was carried out via an incision over the location where a maximum radio-isotype activity could be measured using a mobile manual scintillation probe (C-Trak, Care Wise Medical Products Corporation). Surgery was terminated when no further radioactive foci could be identified in the surgical field. Samples were pseudonymized by the involved medical doctors so that all samples could be processed by non-medical personnel as well.

### Method details

#### Whole-mount procedure staining and tissue clearing

Directly after excision, samples were washed in isotonic sodium chloride solution, photographed and chemically fixed using excess volume of 4% PFA in PBS (CatNo: 9143.1, C. Roth) for 24 h at 4 C. After fixation, SLN were macroscopically cut if the smallest dimension (future z in LSFM) exceeded 6 mm, the working distance of the objective. The other two dimensions did not exceed 13 × 11 mm, avoiding multi-positioning and subsequent stitching algorithms while simultaneously reducing acquisition times. The incisions were made according to routine histopathological guidelines. For tissue permeabilization, samples were incubated at room temperature (RT) in 15 ml tubes (CatNo: 188271, Greiner Bio-one) with permeabilization buffer consisting of 20% DMSO (CatNo: 41641, Fluka), 1% TritonX-100 (CatNo: 3051.3, Carl Roth), 2.3 g glycine/100 ml (CatNo: 3908.2, Carl Roth) in Roti®-CELL PBS (CatNo: 9143.1, Carl Roth). During all incubation steps, samples were shaken in 3-D (Model No: 88881002, Thermo Fisher and RS-DS 5, Phoenix Instrument) except when described otherwise. Overhead rotation was avoided to prevent samples from getting stuck and running dry. Afterwards, samples were dehydrated in an ascending ethanol (EtOH) series, consisting of 50% EtOH (1:1 dilution of 100% EtOH, CatNo: 9065.4, Carl Roth, in ddH_2_O), 70% EtOH (CatNo: T913.2, Carl Roth) and 100% EtOH in 15 ml tubes while at 4°C. Incubation times in 50% EtOH and 70% EtOH were 4 h and 2 h in 100% EtOH.

The bleaching solution was prepared freshly, 10 minutes prior to sample incubation, by diluting 35% H_2_O_2_ (CatNo: 349887, Sigma Aldrich) to 7.5% in precooled bleaching solution, consisting of 5% DMSO in 100% EtOH. After 10 minutes, samples were transferred to these tubes, which were only loosely capped and incubated standing upright for 4 h at 4°C. Then, samples were rehydrated by successive incubation in 70% EtOH and 50% EtOH for at least 4 h at 4°C. While in 50% EtOH, the samples were warmed to RT and washed with 15 ml PBS for 30 minutes. Blocking was conducted for at least 30 minutes while shaking in 2 ml blocking solution containing 0.1% saponine (CatNo: 4185.1, Carl Roth), 0.1% Triton X-100, 0.02% NaN_3_ (CatNo: S2002, Sigma) and 5% donkey serum (CatNo: P30-0101, PAN Biotech) in 1x PBS in amber 2 ml safe-lock tubes (CatNo: 0030120.248, Eppendorf). Incubation was done in a dry incubator (HERAcell 240i, Thermo Fisher) heated to 37 °C. Subsequently, the samples were incubated with fluorophore-conjugated antibodies for another 5 d at 37 °C shaking. For the study, the panel was: 2 µg/ml of anti-CD3 AlexaFluor 647 (CatNo: 344826, Clone: SK7, BioLegend, RRID: AB_2563441) and 10 µg/ml of anti-MelanA (CatNo: NBP2-46603, NovusBio, RRID: AB_2827790) which was self-conjugated with AF 790 (coupling kit: CatNo: A20189, Thermo Fisher) according to the manufacturer’s manual. Note, that the MelanA signal was placed in the channel with the longest possible wavelength to ensure a robust detection of MelanA positive melanoma cells throughout the entire sample, since this was the most important feature for evaluating the clinical applicability of LSFM. In order to be able to compare MelanA positive cells in the sample in LSFM and follow-up FFPE histology, we applied the same antibody clone (A103) used in local clinical routine diagnostics. For the B cell staining, we used 1 µg/ml anti-CD19 AF647 (CatNo: 302220, Clone: HIB19, BioLegend, RRID: AB_389335). After antibody incubation, samples were kept in black 5 ml tubes (CatNo: PE69.1, C. Roth) to protect them from light while being washed twice in 5 ml blocking solution without donkey serum. First, they were incubated for 4 h and then overnight at RT. Samples were then dehydrated in an ascending ethanol series of 50% EtOH, 70% EtOH and 2x 100% EtOH for 4 h at RT in each step. The samples were transferred to brown glass vials (CatNo: 102452 with screw caps CatNo: 102346, Glas-Shop.com) containing pure, fluid ethyl cinnamate (ECi, 99%, CatNo: 112372, Sigma Aldrich) as the refractive index matching agent and incubated for at least 8 h at RT. *Tip: Invert samples at least once 2 h before imaging for optimal clarity*.

#### Light sheet fluorescence microscopy (LSFM)

Samples were imaged using an Ultramicroscope II and ImSpector software (both LaVision BioTec). The microscope is based on a MVX10 zoom body (Olympus) with a 2x objective (NA = 0.5) and equipped with a Neo sCMOS camera (Andor). For image acquisition, samples were either screwed or glued (Loctite 408) on a commercially available sample holder. Beware that contact with glue renders the tissue surface milky. If gluing a sample, it should not be imaged from a different angle afterwards. To avoid damage or deformation of the sample if screwing it tight, ECi-cleared 1% phytagel (CatNo: P8169–100G, Sigma Aldrich) in tap water blocks were used as buffers between tissue and plastic holder, from both sides and from below. Samples were placed on the longest cutting edge, with a perpendicular cutting-edge facing left, if applicable. This allowed correlation with following histopathology. The cleared samples were then immersed in an ECi-filled quartz cuvette and excited with light sheets of different wavelengths (488, 561, 639 and 785 nm). Depending on the fluorophores, following band-pass emission filters were used (mean nm / spread): 525/50 nm or 595/40 nm for tissue autofluorescence; 680/30 nm for AF647 and 835/70 nm for AF790. For tumor screening, the sample was illuminated from both sides with three light sheets from different angles. Optical slices were taken every 10 µm at a low total magnification between 1.26x-2x, providing a detector resolution of 10.32-6.5 µm, respectively. The NA was set to 0.029, yielding a light sheet thickness of approximately 10 µm at the horizontal focus. During overview imaging, samples were evaluated live noting clarity, LN structure and, if applicable, MelanA signal position. Regions of interest, e.g. presumably MelanA positive regions, were subsequently imaged, mostly using a total magnification of 6.4x with a detector resolution of 2.02 µm. Here, acquisition parameters varied depending on size and quality of the region of interest. Sheet width was always at maximum to ensure a homogenous illumination of the sample. Illumination time was set to 350 ms and the total acquisition time of one plane was thus, theoretically, 700 ms. Assuming a z stack as deep as 6 mm, this would have resulted in the acquisition of 600 images × 3 channels = 1800 images, which would theoretically have taken 21 min. However, in our practical experience including loading the samples, the overview imaging of one sample took 30–40 min. Depending on the number of regions of interest, the additional imaging time per sample ranged from 0-2h. To avoid photobleaching during imaging, the longest wavelength was imaged first. The output was a 16bit OME.TIF stack.

For more information on zoom factors, corresponding numerical apertures and resolutions, please visit: https://www.lavisionbiotec.com/products/UltraMicroscope/specification.html.

#### Declarification and gold standard formalin-fixed paraffin-embedded (FFPE) histology

After image acquisition, the samples were declarified in a descending ethanol series of 2x 100% EtOH, 1x 70% EtOH and 1x 50% EtOH for 4 h while shaking at RT. Then, samples were transferred into a plastic cassette (CatNo: M491-5, Simport) and stored in PBS. Subsequently, they were routinely paraffinized using a vacuum infiltration tissue processor (Tissue-Tek VIP 6 AI, Sakura). Paraffin embedding was done using a TBS88 (Medite). Sequential slices of 1.5 µm thickness were acquired by cutting the pre-cooled paraffin blocks (HistoStar cooling plate, Thermo Fisher) using a HM340E microtome (Thermo Fisher) with A35 microtome blades (Feather). Slices were transferred to a water bath set to 40°C (CatNo: MH8517, Electrothermal) and mounted on SuperFrost microscope slides (CatNo: 03-0060, R. Langenbrinck). Slides were stored at RT in the dark, while histopathologically relevant slides according to the local guidelines were routinely processed. H&E staining was done using a Ventana HE 600 (Roche) with Hematoxylin (CatNo: 07024282001) and Eosin (CatNo: 06544304001, Ventana). Immunohistochemistry against MelanA (CatNo: 790-2990) or S100 (CatNo: 790-2914) in red (CatNo: 760-501, all Ventana) was conducted using a BenchMark Ultra (Roche).

#### Routine staging and follow-up histology

Slides were routinely histopathologically assessed by local histopathologists and patients were staged according to these gold standard findings alone. Histopathologists were only informed of patient’s study participation but not about LSFM findings to avoid bias. If live LSFM findings did not match the reported diagnosis, LSFM data were reviewed. If MelanA positivity could be determined in LSFM only, LSFM-guided FFPE follow-up stainings of stored back-up slides were used to confirm the findings. These were then again assessed by histopathologists. An Aperio At2 (Leica) was used to image FFPE slides.

#### In-silico 3-D analysis

16bit OME.TIF stacks were converted (ImarisFileConverterx64, Version 9.3.1, BitPlane) into Imaris files (.ims). 3D reconstruction and subsequent analysis was done using Imaris software (Version 9.3.1, BitPlane). Segmentation of whole tissue including fatty tissue and CD3 positive volume was done using segmentation algorithm on tissue autofluorescence with 20 µm grain size and manual threshold. Segmentations of LN tissue (via LN capsule and/or cutting edges), MelanA signal, blood vessels, B cell follicles and germinal centers are all based on the contour tracing tool and were carried out manually/semi-automatically. For surface creation of the LN tissue every 10^th^ optical slice (100 µm), for all other surfaces every optical slice was traced. Surfaces were created with maximum resolution (2160 × 2560 pixel) and preserved features to ensure matching of the traced lines with the surface border. Discrimination of B cell follicles was based on T cell absence within the LN stroma.

#### Multiscale contrast compression algorithm – RAYhance

For a given mean level of brightness the human visual system (HVS) typically is able to discriminate just about 100 different grey levels (Kalender, 2005; Wandell, 1995). Therefore, we developed RAYhance in order to access the wealth of information hidden in the broad dynamical range of fluorescence microscopy images in one glimpse. It presents slightest contrasts in fine details both in low and high intensity regions simultaneously and keeps strong contrasts under control. Mastering this task provides an image that delivers all structural information at once to the HVS and overcomes the cumbersome and time-consuming task of scanning the entire image by changing the windowing appropriately.

RAYhance modifies the contrast of the input images and its concept of contrast is designed to operate independently of the absolute intensity level of the input image. This allows for processing a whole 3D-stack of LSFM images without generation of artificial discontinuities in image brightness – the whole 3D-volume is processed seamlessly along the direction of the light sheet travelling through the specimen.

Each primary image, in our case uncropped 16bit OME.TIF optical slices, is converted in consecutive steps into several multiscale contrast images that obey the characteristics of the HVS as their contrast is given in units of *just noticeable differences* (JNDs). At each scale-level the JND images undergo non-linear processing of their pixel values that enhances subtle contrasts and decreases high contrasts that are perceivable anyhow. Afterwards, the processed multiscale contrast images are recombined into one grey-level image: the resulting RAYhance-processed image.

RAYhance’s algorithm focuses on content and characteristics of the raw image only and thus runs detector independently. The parameters can be tuned to the specific task and, once set, RAYhance works without any user interaction, giving reproducible results even under different intensities of the illuminating laser.

RAYhance is currently available upon request from LaVision BioTec GmbH. In order to use RAYhance, please send your request to.

#### Shrinkage determination

Generally, samples were photographed macroscopically after excision, fixation, sectioning (if applied), bleaching (after rehydration) and clearing (before LSFM) in order to keep track of the macroscopically visible pigmentation and the shrinkage per sample/piece. However, not all samples were ideally suited for shrinkage measurement of LN tissue due to e.g. fat enrichment. Using FIJI (Schindelin et al., 2012), the area of the samples imaged from directly above was measured using the polygon tracing tool. With the millimeter paper in the background as a reference, the shrinkage per dimension of human LN tissue was determined to be: 5.1% ± 2.4% (arithmetic mean ± SD) from fresh to fixed, 5.9% ± 2.7% from fixed to bleached and another 10.5% ± 1.6% from bleached to cleared. The overall shrinkage from fresh to cleared was 20% ± 5.4% (all N=3). Paraffination of the samples caused further shrinkage compared to the cleared state, which amounted to 18.4% ± 4.6% (N=4).

### Quantification and statistical analysis

In order to compare the volume of primary and secondary follicles in one sample (Figure 4), a two-tailed Mann-Whitney *U*-test was performed. Analysis was performed on all follicles located in a single LN-piece. Results are displayed as median volume ± interquartile range.

## Data Availability

Source data for all figures based in the paper based on Imaris quantification is available [https://doi.org/10.17185/duepublico/71558]. Raw LSFM data is available upon request.

## Data and code availability and additional resources

### Data management and data availability

The data management plan can be accessed via DuEPublico. DOI: 10.17185/duepublico/70526. Source data for all figures based in the paper based on Imaris quantification is available [https://doi.org/10.17185/duepublico/71558]. Raw LSFM data is available upon request.

### Code availability

The Sourcecode of RAYhance cannot be disclosed due to commercial interests. RAYhance is currently available upon request from LaVision BioTec GmbH. In order to use RAYhance, please send your request to.

### Light sheet resources

For more information on the microscope like zoom factors, corresponding numerical apertures and resolutions, please visit: https://www.miltenyibiotec.com/DE-en/products/ultramicroscope-ii.html#scrollto_specs

### Clinical trial registry

The trial description can be accessed under the ID: DRKS00015737 or here: https://www.drks.de/drks_web/

### ECi trouble-shooting FAQ

http://experimental-immunology.de/pages/eci.php

## Acknowledgements

We thank the Imaging Center Essen (IMCES, https://imces.uk-essen.de) for continuous support of our imaging experiments. We also express our thanks for the support by Miltenyi Biotec, LaVision BioTec and the Pathology Unit of the University Hospital Essen. Furthermore, we thank S. Rehwald, S. Hendriks of the University Library Duisburg-Essen and S. Beyer and team of the Centre for Information and Media Services, University Duisburg-Essen for their continuous support concerning data management and IT infrastructure, respectively. This work was supported by the German research Foundation (KFO337 Phenotime to M.G.).

## Author contributions

These authors contributed equally: SFM, PJ

These authors jointly supervised the work: JK, MG

PJ, JK and IS organized the clinical study, recruited patients and performed surgery. SFM and PJ developed and implemented the workflow and carried out the study supported by RU, AK and LB. SFM did LSFM imaging and in-silico processing, assisted by ZC, LB, RU, PJ. RU was responsible for FFPE sectioning. EH, KG and TS assessed the samples histopathologically. RAYhance/FullView was developed by GE and TA. MG, JK, IS, SB, DS and EH provided supervision. JK, MG and IS conceived of the study and wrote the manuscript together with SFM, PJ, EH, GE and KG. All authors contributed to discussion and correction of the manuscript.

## Supplementary Materials

Supplementary Figure 1: Algorithm-enhanced LSFM autofluorescence signal for anatomical sample overview

Supplementary Figure 2: RAYhance – a multiscale compression algorithm for simultaneous display of LSFM data with high signal intensity spread Supplementary materials and methods

Supplementary Figure 3: LSFM analysis allows subsequent immunohistochemical formalin-fixed paraffin-embedded (FFPE) gold standard stainings

Supplementary Figure 4: Unspecific cutting-edge MelanA artifact

Supplementary Figure 5: Infrared (IR) dye enables imaging of tumor through residual pigmentation

Supplementary Table 1: Patient characteristics

Supplementary Table 2: Tissue statistics and diagnostic results per patient

Supplementary Table 3: Sample preparation time of SLN analysis and LSFM module for melanoma diagnostics

Supplementary Video 1: An entire human LN in 3-D based on autofluorescence Supplementary Video 2: Interplay of melanoma and T cells in a human LN (hLN) Supplementary Video 3: RAYhance for browsable histo-like data stacks Supplementary Video 4: From 2-D to 3-D – a capsular nevus

Supplementary Video 5: From 3-D to quantification – segmentation of a capsular nevus Supplementary Video 6: Anatomical and cellular LN architecture 3-D quantified Supplementary Video 7: 3-D quantification of melanoma

Supplementary Video 8: Tumor detection through pigmentation Supplementary Video 9: LSFM detects “hidden” metastasis

Supplementary Note 1: LSFM artifacts and their interpretation – concerning tumor detection through residual pigmentation

